# Mendelian randomization analysis to characterize causal association between coronary artery disease and COVID-19

**DOI:** 10.1101/2020.05.29.20117309

**Authors:** Lang Wu, Jingjing Zhu, Chong Wu

## Abstract

Observational studies have suggested that having coronary artery disease increases the risk of Coronavirus disease 2019 (COVID-19) susceptibility and severity, but it remains unclear if this association is causal. Inferring causation is critical to facilitate the development of appropriate policies and/or individual decisions to reduce the incidence and burden of COVID-19. We applied Two-sample Mendelian randomization analysis and found that genetically predicted CAD was significantly associated with higher risk of COVID-19: the odds ratio was 1.29 (95% confidence interval 1.11 to 1.49; *P* = 0.001) per unit higher log odds of having CAD.

## Introduction

Coronavirus disease 2019 (COVID-19) has become a global pandemic. Specific risk factors such as coronary artery disease (CAD) ^1^ have been reported to be related to COVID-19 susceptibility and severity through conventional observational studies. However, findings from conventional observational studies are susceptible to selection bias, unmeasured confounding, and reverse causation. With these limitations, it is very challenging to establish the causality of reported associations in observational studies. Inferring causation is critical to facilitate the development of appropriate policies and/or individual decisions to reduce the incidence and burden of COVID-19. Here we show that the Mendelian Randomization (MR) design, similar to a “genetic randomized controlled trial”, can be adopted to decipher the causality of the association between CAD and COVID-19 ^2^.

## Methods

### Genetic association datasets for COVID-19 susceptibility

For evaluation of associations with COVID-19 risk, we used summary statistics data of the most recent version of genome-wide association study (GWAS) analyses from The COVID-19 host genetics initiative (released on May 17, 2020) ^3^. Detailed information on participating studies, quality control and analyses has been provided on the COVID-19 host genetics initiative website (https://www.covid19hg.org/results/). In brief, data from approximately 1,678 affected cases and 674,635 uninfected controls from studies of BioMe, FinnGen, Genes & Health, LifeLines Global Screening Array, LifeLines CytoSNP, Netherlands Twin Register, Partners Healthcare Biobank, and UK Biobank were used. The majority of the included subjects are Europeans, with a small proportion of other ethnic groups. Only variants with imputation quality > 0.6 were retained. Meta-analysis of individual studies was performed with inverse variance weighting.

### Instrumental variables for CAD risk

We extracted genetic instruments for CAD risk based on a comprehensive GWAS of ∼185,000 CAD cases and controls by Nikpay et al ^4^. In this study, a majority of the participants (77%) were of European ancestry, and 19% were of Asian ancestry. We selected single nucleotide polymorphisms (SNPs) associated at *P*< 5×10^−8^ and only retained independent variants from each other (r^2^ < 0.001). For correlated SNPs, the SNP with the lowest *p*-value was selected.

### MR analysis

We applied the widely-used inverse variance weighted (IVW) method ^5^ to estimate the overall causal association of CAD on COVID-19 susceptibility. To account for potential violations of valid instrumental variable assumptions, we conducted sensitive analyses by applying several methods that are robust to horizontal pleiotropy at the cost of reduced statistical power. These include weighted median MR, MR-Egger regression, and MR-PRESSO test, and leave-one-out analysis. Results are presented as odds ratios per unit higher log odds of CAD.

## Results

For MR analysis, we used 38 independent variants as the instrument for CAD (data not shown) with an estimated F-statistic of 62. Genetically predicted CAD was significantly associated with higher risk of COVID-19. The odds ratio was 1.29 (95% confidence interval 1.11 to 1.49; *P* = 0.001) per unit higher log odds of having CAD (**Figure 1**). The association remained robust in weighted median MR (**Figure 1**) and leave one out analyses (**Figure 2**). There was limited evidence of horizontal pleiotropy and heterogeneity based on the MR-Egger intercept test (*P* = 0.83) and MR-PRESSO (*P* = 0.26).

**Figure 1.**
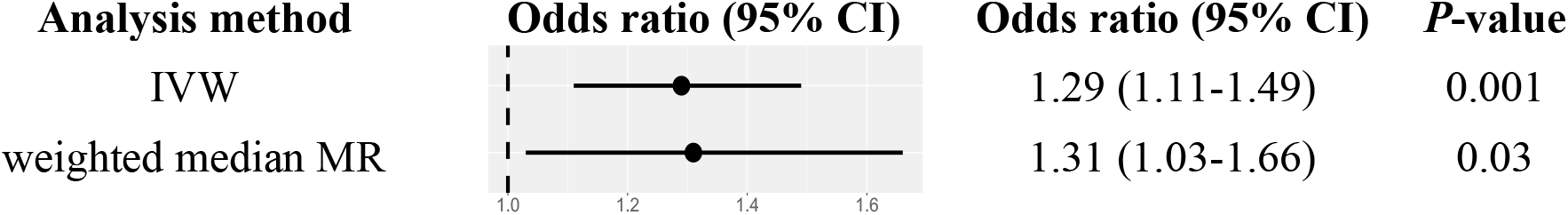
Odds ratios for associations between genetically predicted higher log odds ratio of having CAD and COVID-19 susceptibility. IVW represents the widely-used inverse variance weighted method and weighted median MR is a robust MR method that provides a consistent estimator when up to 50% of the instrumental variables are invalid.

**Figure 2.**
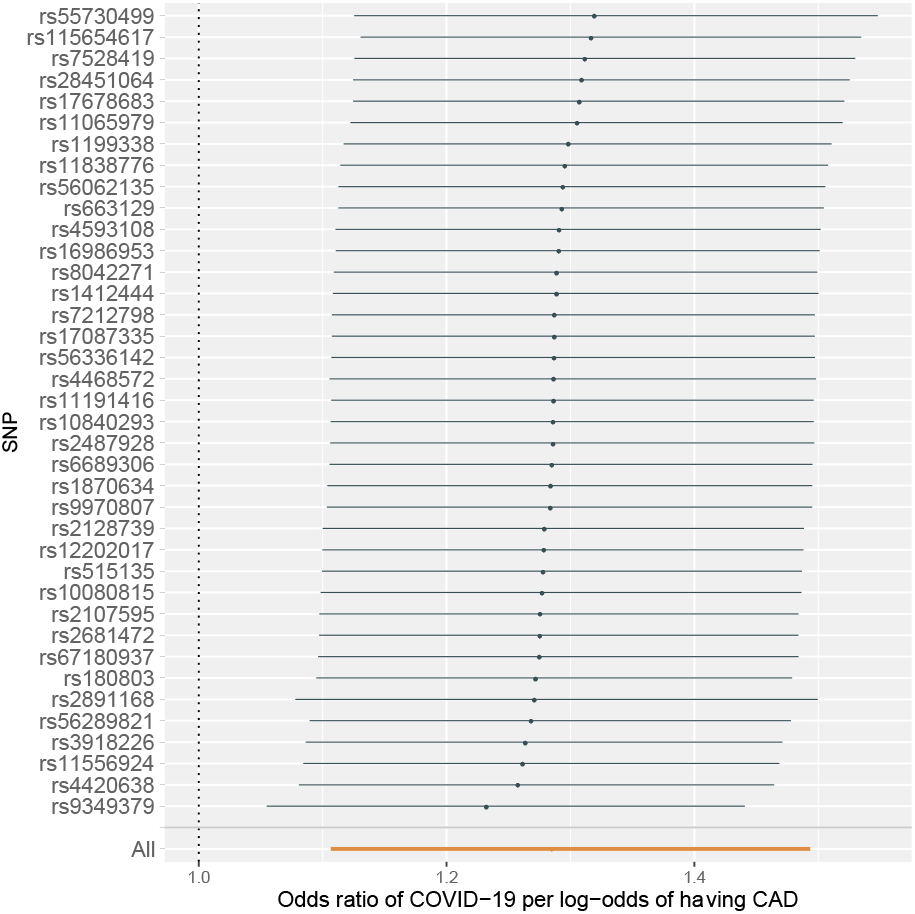
Leave-one-out analysis: each row represents a MR analysis of CAD on COV-19 using all instruments expect for the SNP listed on the y-axis. The point represents the odds ratio with that SNP removed and the line represents 95% confidence interval.

## Discussion

In an MR analysis, we observed evidence supporting a potential causal association between CAD and COVID-19. Such a link may be explained by changes of viscosity during febrile illnesses, heightened coagulation systems, endothelial cell dysfunction, or proinflammatory effects ^6^. This is the first study to characterize potential causality of suspected risk factors for COVID-19 susceptibility using a MR design. Limitations of the current study include 1) the overlap of relatively small samples in both GWAS of CAD and COVID-19; and 2) mixed population composition in both GWAS of CAD and COVID-19. Because BioMe only contributed a relatively small sample size to the COVID-19 GWAS: 20 (1.1%) cases and 10,169 (1.5%) controls, the potential influence of this on the MR estimate should be small. In both GWAS of CAD and COVID-19, a majority of the subjects are Europeans. In summary, an MR study could potentially avoid many biases and confounding issues existing in conventional observational studies and thus help to identify causally related risk factors. Using MR design, we found evidence that having CAD is associated with a higher risk of COVID-19. Therefore, particular attention should be given to individuals with CAD during this pandemic and we expect that this finding will facilitate appropriate policymaking and individual decisions to reduce COVID-19 burden.

## Data Availability

We used the following two public available GWAS summary data for our analysis: 1. data from the COVID-19 Host Genetics Initiative (https://www.covid19hg.org); 2. data from IEU GWAS database (https://gwas.mrcieu.ac.uk).

